# Common and Rare Variants Associated with Cardiometabolic Traits across 98,622 Whole-Genome Sequences in the *All of Us* Research Program

**DOI:** 10.1101/2022.11.23.22282687

**Authors:** Xin Wang, Justine Ryu, Jihoon Kim, Andrea Ramirez, Kelsey R. Mayo, All of Us Research Program, Henry Condon, Nataraja Sarma Vaitinadin, Lucila Ohno-Machado, Greg A. Talavera, Patrick T. Ellinor, Steven A. Lubitz, Seung Hoan Choi

**Affiliations:** Cardiovascular Research Center, Massachusetts General Hospital, Boston, Massachusetts, USA; Cardiovascular Disease Initiative, The Broad Institute of MIT and Harvard, Cambridge, Massachusetts, USA; Department of Biomedical Informatics, University of California San Diego Health System, La Jolla, California, USA; All of Us Research Program, National Institutes of Health, Bethesda, Maryland, USA; Vanderbilt Institute of Clinical and Translational Research, Vanderbilt University Medical Center, Nashville, Tennessee, USA; Department of Genetic Medicine, Vanderbilt University Medical Center, Nashville, Tennessee, USA; Department of Medicine, Vanderbilt University Medical Center, Nashville, Tennessee, USA; Graduate School of Public Health, San Diego State University, San Diego, California, USA; Demoulas Center for Cardiac Arrhythmias, Massachusetts General Hospital, Boston, Massachusetts, USA; Department of Biostatistics, Boston University, Boston, Massachusetts, USA

## Abstract

*All of Us* is a biorepository aiming to advance biomedical research by providing various types of data in diverse human populations. Here we present a demonstration project validating the program’s genomic data in 98,622 participants. We sought to replicate known genetic associations for three diseases (atrial fibrillation [AF], coronary artery disease, type 2 diabetes [T2D]) and two quantitative traits (height and low-density lipoprotein [LDL]) by conducting common and rare variant analyses. We identified one known risk locus for AF, five loci for T2D, 143 loci for height, and nine loci for LDL. In gene-based burden tests for rare loss-of-function variants, we replicated associations between *TTN* and AF, *GIGYF1* and T2D, *ADAMTS17, ACAN, NPR2* and height, *APOB, LDLR, PCSK9* and LDL. Our results are consistent with previous literature, indicating that the *All of Us* program is a reliable resource for advancing the understanding of complex diseases in diverse human populations.

## Main text

The *All of Us* Research Program (*All of Us*) is a prospective cohort study launched in 2018 with the goal of improving population-based research and advancing understanding of human disease. To this end, *All of U*s plans to enroll at least 1 million individuals living in the United States and collect large-scale electronic health record (EHR) data, laboratory and physical measurements, survey responses, and genomic data.^1^ As of March 2022, *All of Us* has released whole-genome sequencing data for 98,622 participants and genotype array data for 165,208 participants. All enrolled individuals have provided written informed consent to the program. In order to validate the quality of the genomic data, *All of Us* launched demonstration projects aimed at replicating well-established genetic findings within the *All of Us* dataset. Approval to use the dataset for the specified demonstration projects was obtained from the *All of Us* Institutional Review Board.

To date, large-scale genome-wide association studies (GWAS) have identified hundreds of risk loci across the human genome for cardiometabolic traits.^2–6^ In the present study, we analyzed common and rare variants from whole-genome sequencing data in the C2021Q3R6 database version of 98,622 participants. The goal of the current project was to ensure the validity of the *All of Us* dataset, by replicating established genetic associations for five cardiometabolic traits, including atrial fibrillation (AF), coronary artery disease (CAD), type 2 diabetes (T2D), height, and low-density lipoprotein (LDL).

We modified a previously described phenotype algorithm^7^ comprising International Classification of Diseases (ICD) codes, self-reported personal medical history, and procedure and operation codes to define AF in the *All of Us* dataset. For CAD and T2D, we used published phenotype algorithms obtained from the Electronic Medical Records and Genomics (eMERGE) network, which have been implemented in *All of Us’* phenotype library.^8,9^ Height and LDL were extracted from the program’s physical measurements and EHR data, respectively. Detailed phenotyping strategies were included in **Supplemental Methods** and **Table S1**. After removing participants who did not pass the sample quality control (QC) procedures (**Supplemental Methods**), we identified 98,564 participants with a mean age of 51.31 (SD 16.87) at enrollment. Of those, 38,263 (38.82%) were male, and 50,213 (50.94%) were not genetically determined to be of European descent. Characteristics of participants are presented in **Table 1**. After applying the phenotype algorithms to the *All of Us* database, we defined 5,120 (5.19%) AF, 3,544 (3.60%) CAD, and 8,557 (8.68%) T2D patients. Furthermore, we identified 34,538 (35.04%) samples with LDL measurements ascertained from the EHR and 94,842 (96.22%) samples with the *All of Us* measured height available. Using these data, we tested associations between genetic variants and phenotypes, and compared our results to previously published GWAS by estimating genetic correlation.

**Table 1.**
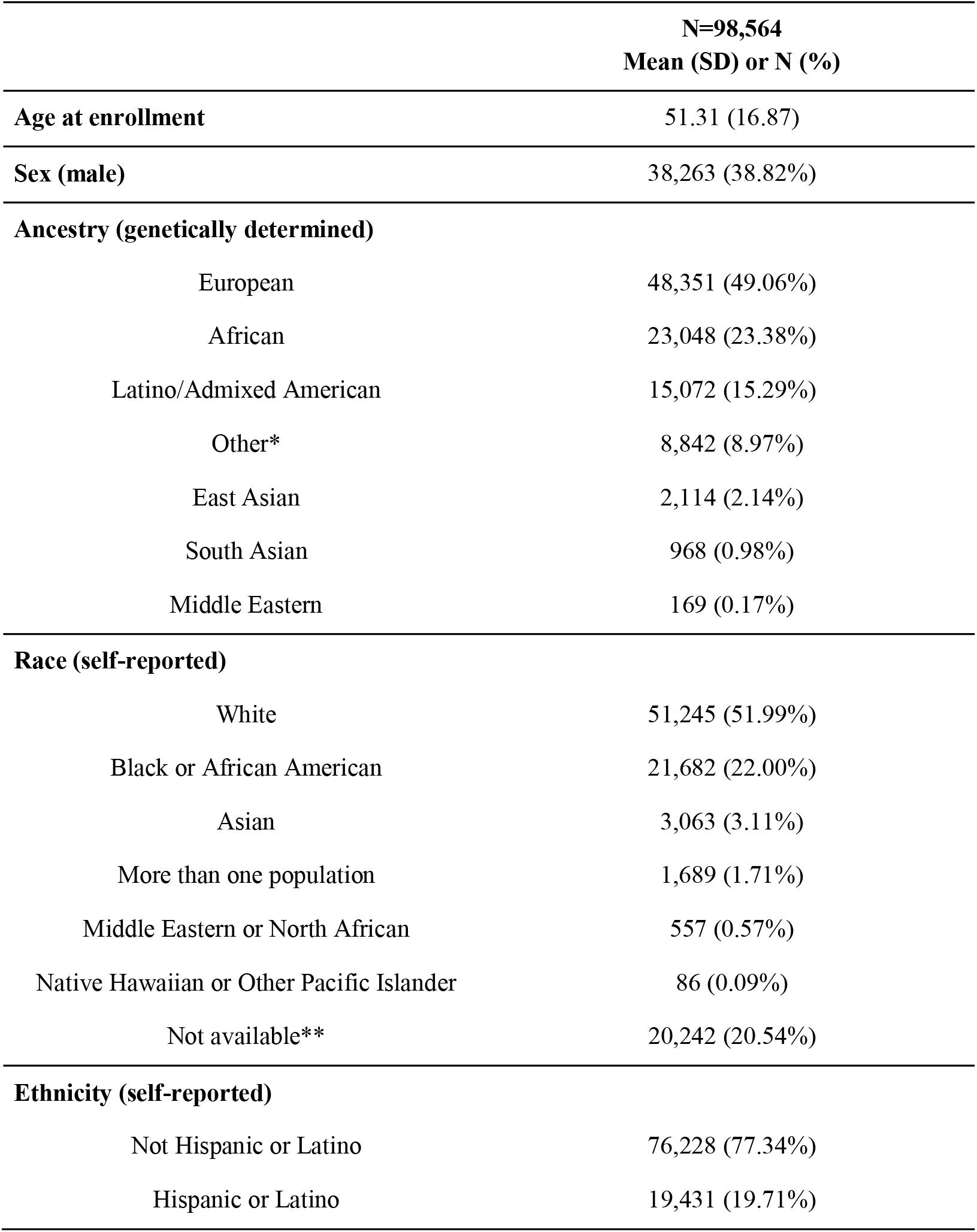
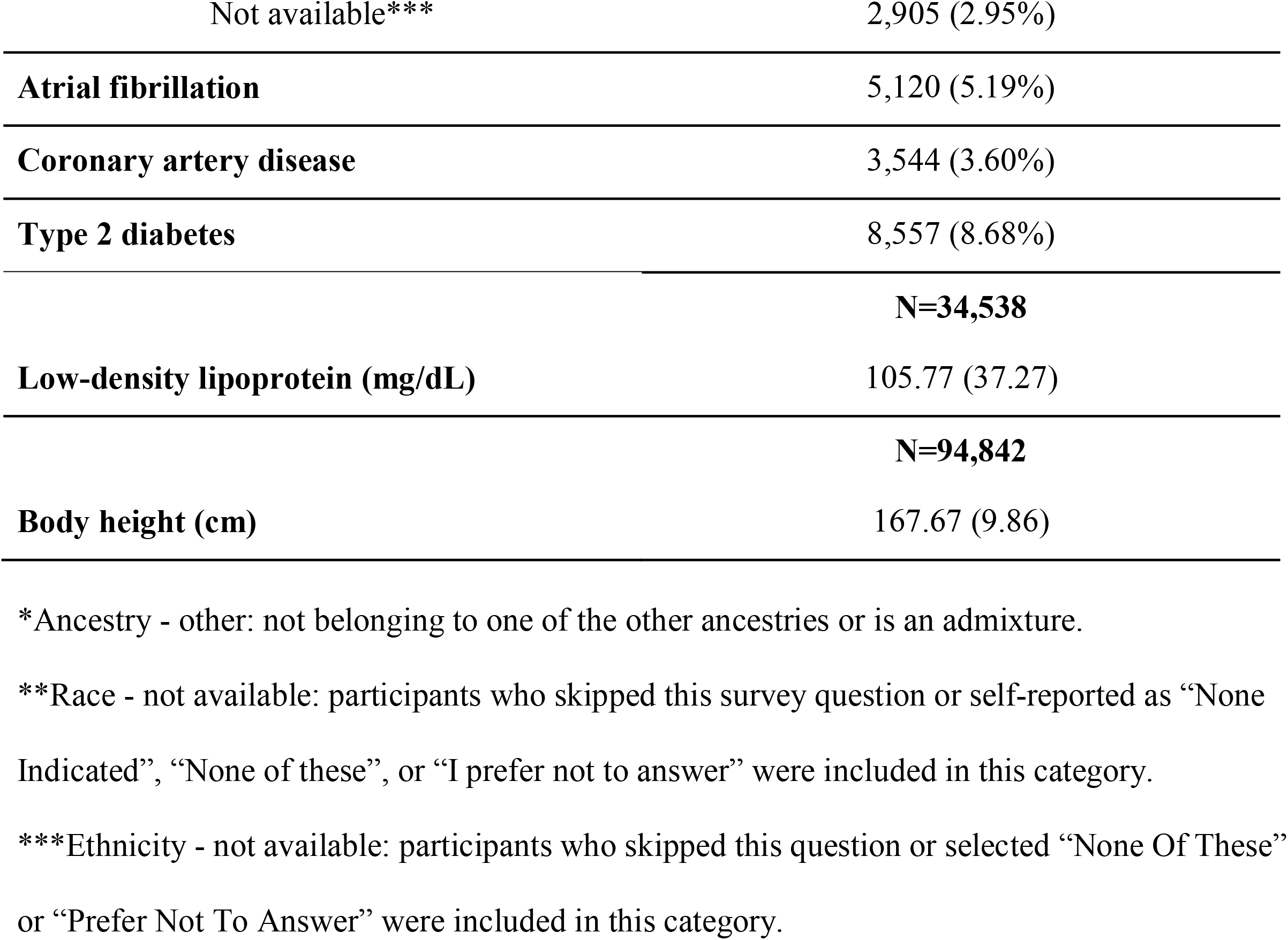
Characteristics of participants in the present study.

|  | **N=98,564**<br>**Mean (SD) or N (%)** |
| --- | --- |
| **Age at enrollment** | 51.31 (16.87) |
| **Sex (male)** | 38,263 (38.82%) |
| **Ancestry (genetically determined)** |  |
| European | 48,351 (49.06%) |
| African | 23,048 (23.38%) |
| Latino/Admixed American | 15,072 (15.29%) |
| Other* | 8,842 (8.97%) |
| East Asian | 2,114 (2.14%) |
| South Asian | 968 (0.98%) |
| Middle Eastern | 169 (0.17%) |
| **Race (self-reported)** |  |
| White | 51,245 (51.99%) |
| Black or African American | 21,682 (22.00%) |
| Asian | 3,063 (3.11%) |
| More than one population | 1,689 (1.71%) |
| Middle Eastern or North African | 557 (0.57%) |
| Native Hawaiian or Other Pacific Islander | 86 (0.09%) |
| Not available** | 20,242 (20.54%) |
| **Ethnicity (self-reported)** |  |
| Not Hispanic or Latino | 76,228 (77.34%) |
| Hispanic or Latino | 19,431 (19.71%) |
| Not available*** | 2,905 (2.95%) |
| **Atrial fibrillation** | 5,120 (5.19%) |
| **Coronary artery disease** | 3,544 (3.60%) |
| **Type 2 diabetes** | 8,557 (8.68%) |
|  | **N=34,538** |
| **Low-density lipoprotein (mg/dL)** | 105.77 (37.27) |
|  | **N=94,842** |
| **Body height (cm)** | 167.67 (9.86) |

\*Ancestry - other: not belonging to one of the other ancestries or is an admixture.
\*\*Race - not available: participants who skipped this survey question or self-reported as “None Indicated”, “None of these”, or “I prefer not to answer” were included in this category.
\*\*\*Ethnicity - not available: participants who skipped this question or selected “None Of These” or “Prefer Not To Answer” were included in this category.

For common variants (minor allele frequency [MAF] > 1%), we used a whole-genome regression approach implemented in the REGENIE^10^ software to test the association between each phenotype and individual single nucleotide variants (SNVs) assuming an additive genetic model, adjusting for age (enrollment age for disease phenotypes, measurement age for continuous traits), sex, and top 20 principal components of ancestry. For binary traits, we also accounted for case-control imbalance using the saddle point approximation (SPA) method^11^ implemented in REGENIE. We identified one genome-wide significant (*P* < 5×10^−8^) risk locus (defined as 500kb upstream and downstream of the lead SNV) for AF (**Table 2, Figure 1a**, and **Figure S2**) upstream of *PITX2*, an established susceptibility locus for AF.^2^ Pitx2 is critical for specification of cardiac symmetry, myocardial sleeve development in the pulmonary veins, and suppression of a default sinus node in the left atrium.^12,13^ We did not observe any inflation in the present AF GWAS (genomic inflation factor [λ_gc_]=1.05, LDSC intercept=1.03, [s.e. 0.01]). The genetic correlation between the *All of Us* GWAS and a prior AF GWAS^14^ was 1.02 (s.e. 0.29), estimated using LD score regression (LDSC).^15^ No genome-wide significant signals were identified for CAD (**Figure S1**). We, however, noted that the most significant locus was at chromosome 9 (lead SNV=rs10811656, OR=1.14 [1.08-1.19], *P*-value=6.48×10^−7^) near *CDKN2B-AS1*, which has been reported to be associated with CAD in prior studies.^16^ The *All of Us* CAD GWAS did not demonstrate any inflation (λ_gc_=1.04, LDSC intercept=1.03 [s.e. 0.01]) and has a genetic correlation of 0.89 (s.e. 0.39) with a previous CAD GWAS.^16^ For T2D, we identified five genome-wide significant loci (**Table 2, Figure 1b**, and **Figure S3**) near *HFE, CDKN2B, TCF7L2, CCND2*, and *FTO. HFE* has been linked to glycated hemoglobin (HbA_1c_) levels in a previous report.^17^ *CDKN2B, TCF7L2, CCND2*, and *FTO* have been reported to be associated with T2D.^18–20^ Minimal genomic inflation was observed (λ_gc_=1.11) in the current GWAS, which was likely due to polygenicity rather than population stratification, as indicated by its LDSC intercept (1.03, s.e. 0.01). The genetic correlation between the *All of Us* GWAS and a prior T2D GWAS^18^ was 0.77 (s.e. 0.10).

**Table 2.**
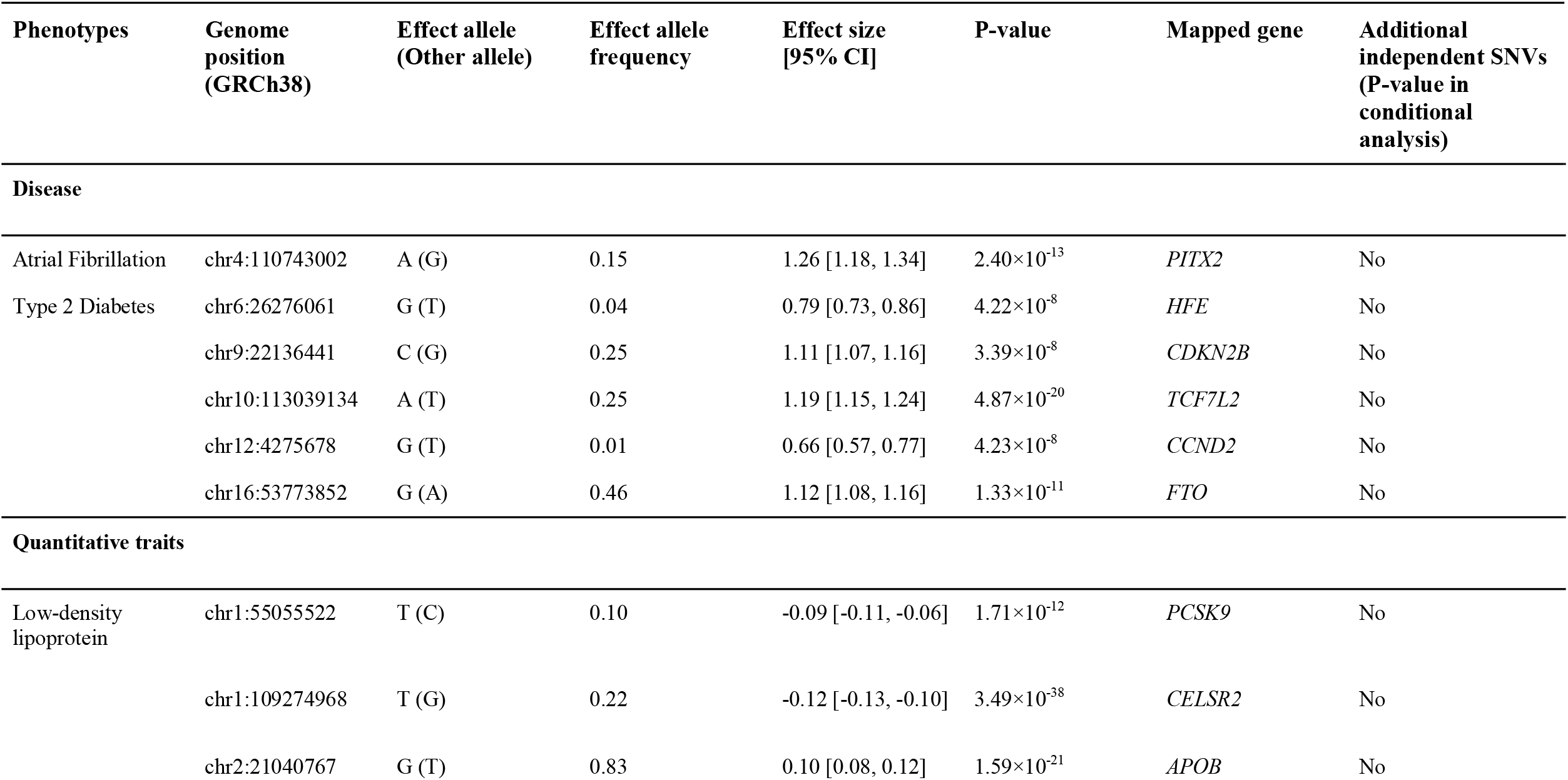
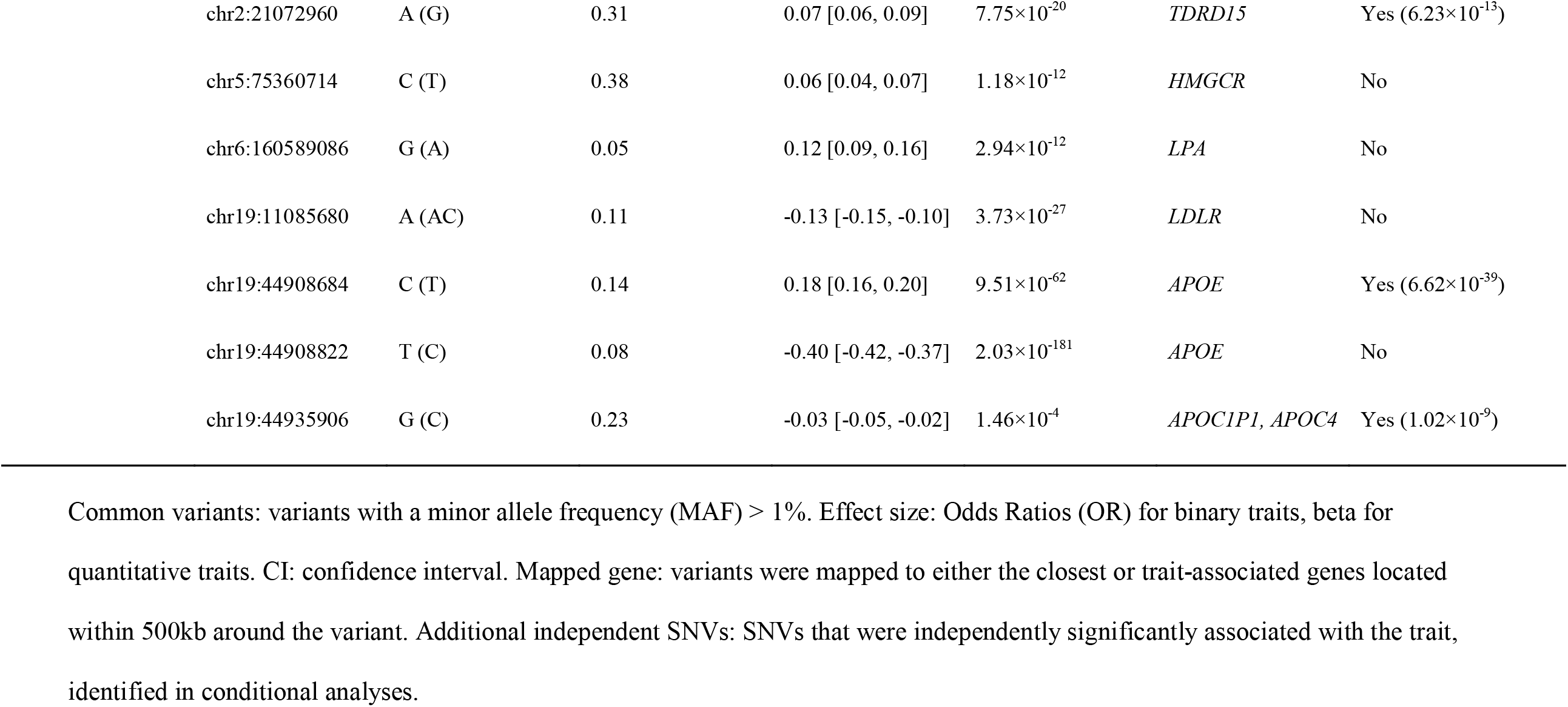
Top associations between phenotypes and common variants.

| Phenotypes | Genome position (GRCh38) | Effect allele (Other allele) | Effect allele frequency | Effect size [95% CI] | P-value | Mapped gene | Additional independent SNVs (P-value in conditional analysis) |
| --- | --- | --- | --- | --- | --- | --- | --- |
| **Disease** |  |  |  |  |  |  |  |
| Atrial Fibrillation | chr4:110743002 | A (G) | 0.15 | 1.26 [1.18, 1.34] | $2.40 \times 10^{-13}$ | *PITX2* | No |
| Type 2 Diabetes | chr6:26276061 | G (T) | 0.04 | 0.79 [0.73, 0.86] | $4.22 \times 10^{-8}$ | *HFE* | No |
| | chr9:22136441 | C (G) | 0.25 | 1.11 [1.07, 1.16] | $3.39 \times 10^{-8}$ | *CDKN2B* | No |
| | chr10:113039134 | A (T) | 0.25 | 1.19 [1.15, 1.24] | $4.87 \times 10^{-20}$ | *TCF7L2* | No |
| | chr12:4275678 | G (T) | 0.01 | 0.66 [0.57, 0.77] | $4.23 \times 10^{-8}$ | *CCND2* | No |
| | chr16:53773852 | G (A) | 0.46 | 1.12 [1.08, 1.16] | $1.33 \times 10^{-11}$ | *FTO* | No |
| **Quantitative traits** |  |  |  |  |  |  |  |
| Low-density lipoprotein | chr1:55055522 | T (C) | 0.10 | -0.09 [-0.11, -0.06] | $1.71 \times 10^{-12}$ | *PCSK9* | No |
| | chr1:109274968 | T (G) | 0.22 | -0.12 [-0.13, -0.10] | $3.49 \times 10^{-38}$ | *CELSR2* | No |
| | chr2:21040767 | G (T) | 0.83 | 0.10 [0.08, 0.12] | $1.59 \times 10^{-21}$ | *APOB* | No |

|  |  |  |  |  |  |  |
| --- | --- | --- | --- | --- | --- | --- |
| chr2:21072960 | A (G) | 0.31 | 0.07 [0.06, 0.09] | $7.75 \times 10^{-20}$ | *TDRD15* | Yes ($6.23 \times 10^{-13}$) |
| chr5:75360714 | C (T) | 0.38 | 0.06 [0.04, 0.07] | $1.18 \times 10^{-12}$ | *HMGCR* | No |
| chr6:160589086 | G (A) | 0.05 | 0.12 [0.09, 0.16] | $2.94 \times 10^{-12}$ | *LPA* | No |
| chr19:11085680 | A (AC) | 0.11 | -0.13 [-0.15, -0.10] | $3.73 \times 10^{-27}$ | *LDLR* | No |
| chr19:44908684 | C (T) | 0.14 | 0.18 [0.16, 0.20] | $9.51 \times 10^{-62}$ | *APOE* | Yes ($6.62 \times 10^{-39}$) |
| chr19:44908822 | T (C) | 0.08 | -0.40 [-0.42, -0.37] | $2.03 \times 10^{-181}$ | *APOE* | No |
| chr19:44935906 | G (C) | 0.23 | -0.03 [-0.05, -0.02] | $1.46 \times 10^{-4}$ | *APOC1P1, APOC4* | Yes ($1.02 \times 10^{-9}$) |

Common variants: variants with a minor allele frequency (MAF) > 1%. Effect size: Odds Ratios (OR) for binary traits, beta for quantitative traits. CI: confidence interval. Mapped gene: variants were mapped to either the closest or trait-associated genes located within 500kb around the variant. Additional independent SNVs: SNVs that were independently significantly associated with the trait, identified in conditional analyses.

**Figure 1.**
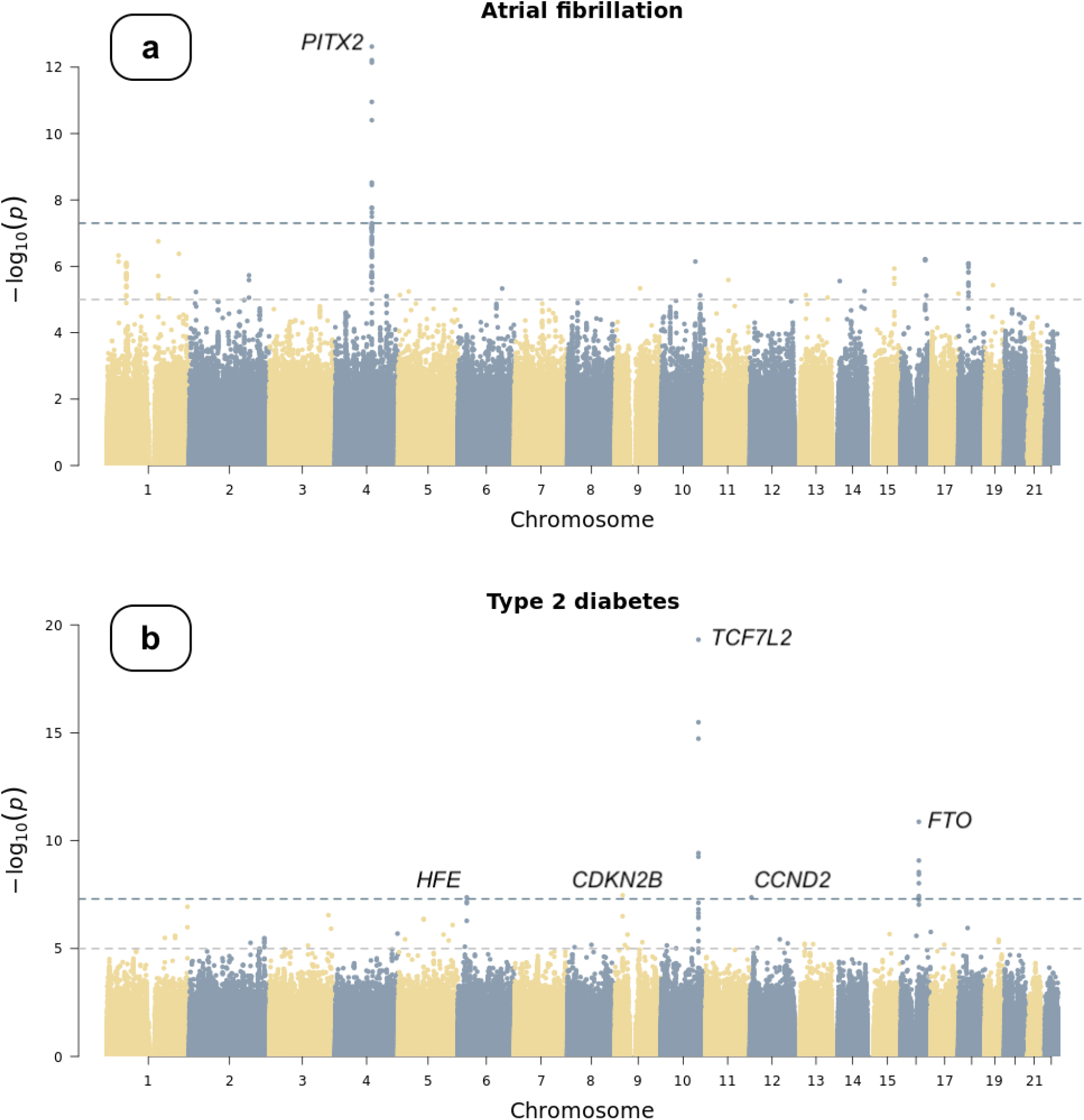
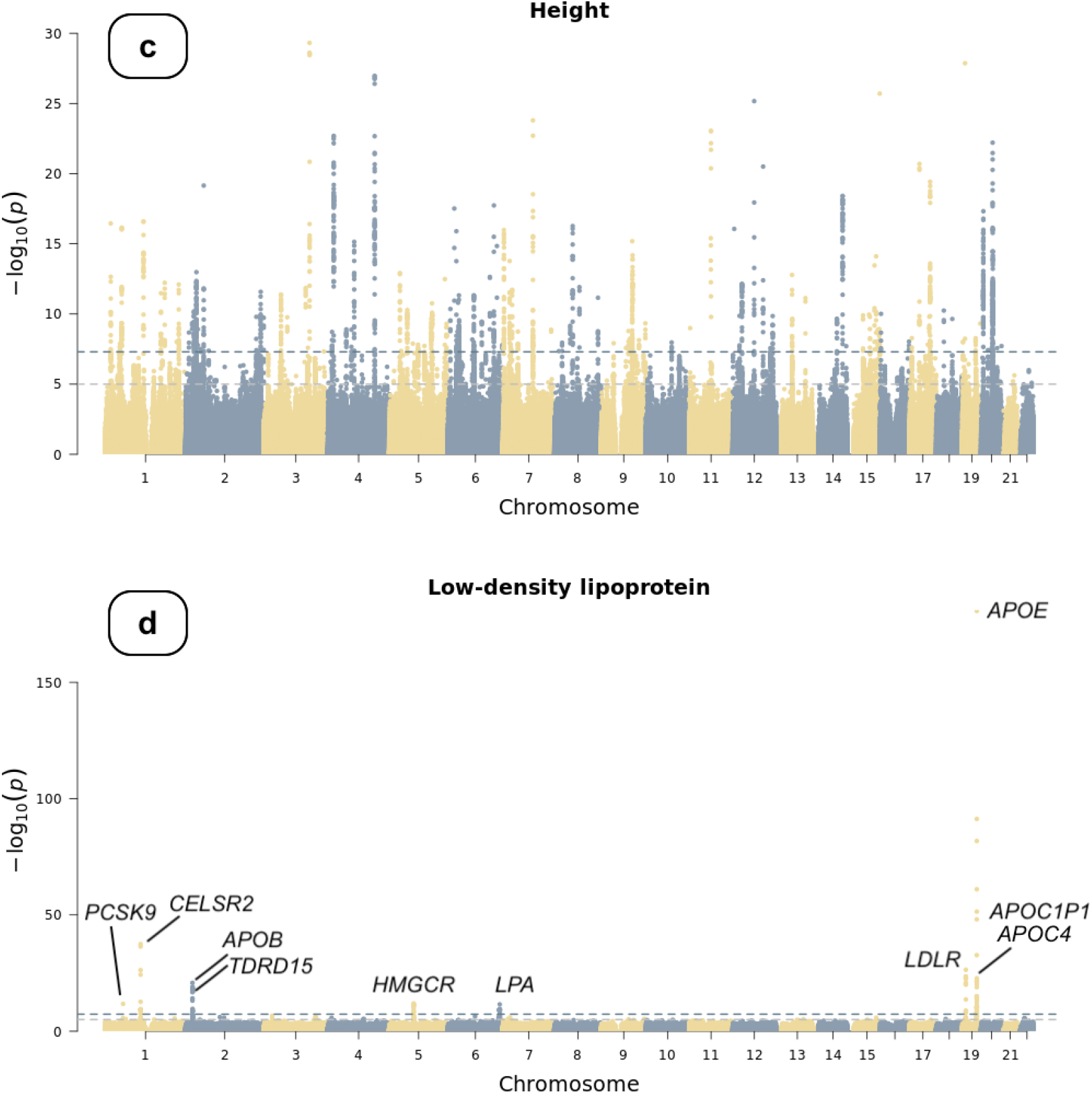
Manhattan plots of genome-wide association studies. Chromosomal variant positions are plotted on the x-axis. The -log10(*P* values) are plotted on the y-axis. The genome-wide significance threshold (5×10−8) is indicated by the horizontal dotted line. Panels display associations with (a) atrial fibrillation, (b) type 2 diabetes, (c) height, and (d) low-density lipoprotein (LDL). Height and LDL were rank-based inverse normal transformed prior to association testing (see text).

We applied rank-based inverse normal transformation (INT) to body height and LDL cholesterol prior to association testing. Using data from 94,842 participants, we identified 143 genome-wide significant loci in the height GWAS. Additionally, we identified 10 secondary independent SNVs at these loci in a conditional analysis using the GCTA software^21^ (**Figure 1c**). The genetic correlation between the *All of Us* GWAS and a previously published height GWAS^6^ was 0.96 (s.e. 0.02). Although the genomic inflation factor of the *All of Us* GWAS was relatively high (λ_gc_=1.33), it was likely due to polygenicity as implicated by its LDSC intercept (1.06, s.e. 0.02) and as height is a highly polygenic trait.^22^ For LDL cholesterol, we identified seven genome-wide significant loci (**Table 2, Figure 1d**, and **Figure S4**) in 34,538 participants implicating *PCSK9, CELSR2, APOB, HMGCR, LPA, LDLR*, and *APOE* gene regions. Three additional independent SNVs were identified in a conditional analysis, implicating *TDRD15, APOE*, and *APOC1P1*/*APOC4* (**Table 2**). Genetic variants at the gene encoding proprotein convertase subtilisin/kexin type 9 (*PCSK9*), the gene encoding cadherin EGF LAG seven-pass G-type receptor 2 (*CELSR2*), the apolipoprotein B and E genes (*APOB* and *APOE*), the gene encoding 3-Hydroxy-3-Methylglutaryl-CoA reductase (*HMGCR*), the LDL receptor gene (*LDLR*), and the gene encoding tudor domain containing 15 (*TDRD15*) have been consistently associated with LDL cholesterol levels.^23–26^ We did not observe any inflation in the current GWAS (λ_gc_=1.02, LDSC intercept=1.01 [s.e. 0.01]). The genetic correlation between the *All of Us* LDL GWAS and a prior GWAS of LDL^27^ was 0.99 (s.e. 0.26). We compared the summary statistics of the lead common genetic variants identified in the present study to those from the corresponding reference GWAS in **Table S2**.

We then sought to replicate known rare variant (MAF < 0.1% and Population Maximum MAF < 0.1%) associations, including those between *TTN* and AF, *LPL* and CAD, *GIGYF1* and T2D, *ADAMTS10, ADAMTS17, ACAN, NPR2* and height, *APOB, LDLR, PCSK9* and LDL.^28^ We first counted the number of sequenced participants who were carriers of high-confidence loss-of-function (LoF) variants of these genes. LoF variants predicted by Loss-of-Function Transcript Effect Estimator (LOFTEE)^29^ can disrupt the function of protein-coding genes and thus may have functional impacts on phenotypes that are associated with these genes. Genes with < 20 carriers were removed from the analysis per *All of Us’* Data and Dissemination Policy [https://www.researchallofus.org/data-tools/data-access/]. We performed gene-based burden tests as implemented in REGENIE,^10^ adjusting for age (enrollment age for disease phenotypes, measurement age for continuous traits), sex, top 20 principal components of ancestry, and case-control imbalance using the SPA method for binary traits. We observed significant associations in the *All of Us* data release for each of the previously reported genes and phenotypes (**Figure 2**). For example, 1 unit increase in the burden of LoF variants within *TTN* was associated with 2.23 [1.65, 2.72] (*P*-value=5.05×10^−8^) times odds of diagnosing with AF. Likewise, 1 unit increase in the burden of LoF variants within *GIGYF1* was associated with 9.03 [3.32, 24.53] (*P*-value=2.39×10^−5^) times odds of diagnosing with T2D, and 1 unit increase in the burden of LoF variants within *APOB* was associated with 1.55 [1.23, 1.90] unit decrease in the INT transformed LDL level. These associations showed the same directional effects and similar effect sizes compared to results from a recently published study using whole-exome sequencing data in the UK Biobank^28^ (**Figure 2)**.

**Figure 2.**
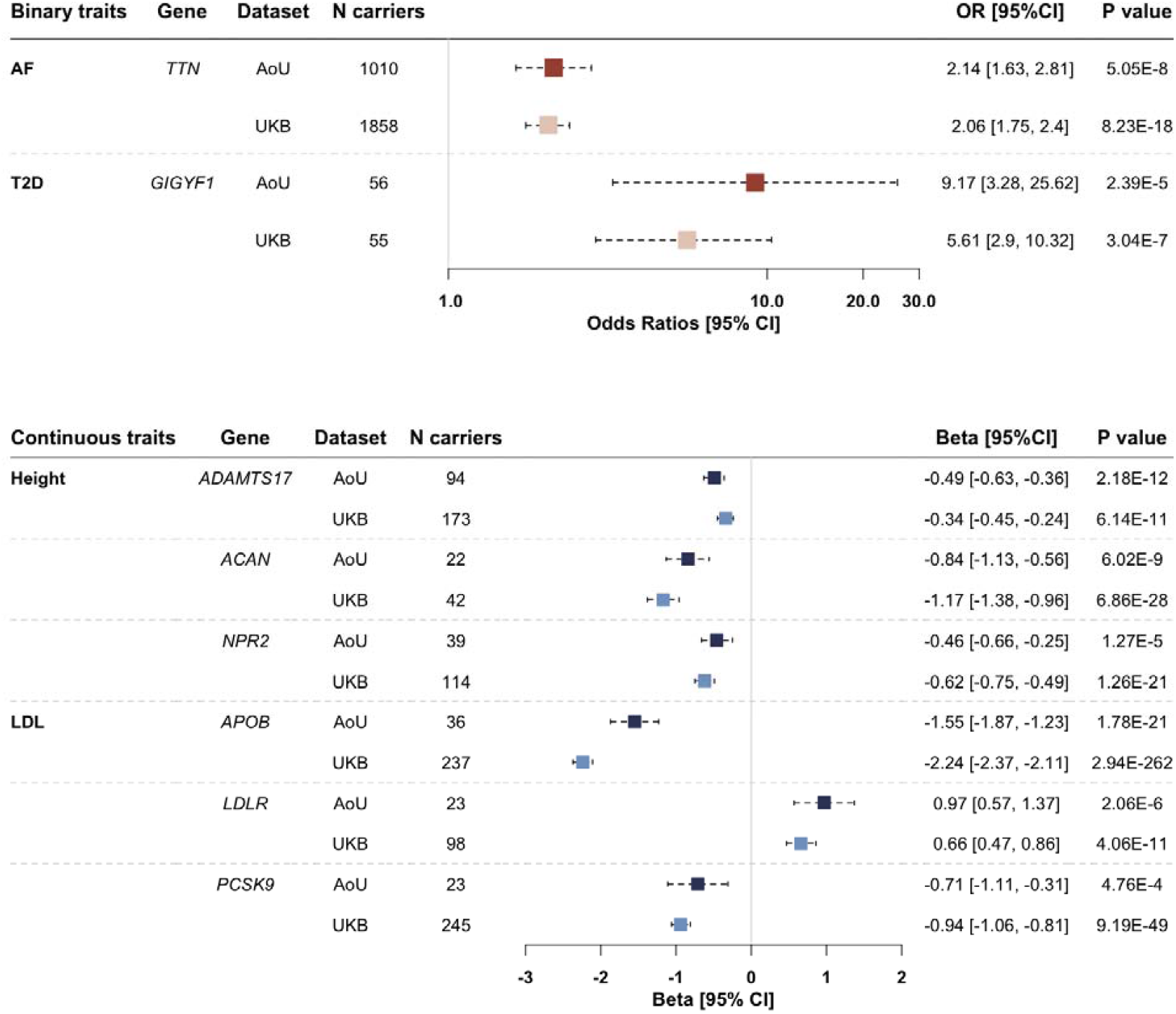
Associations between phenotypes and genes harboring rare variants. Rare variants: rare (minor allele frequency [MAF] < 0.1% and Population Maximum MAF < 0.1%) loss-of-function variants. AF: atrial fibrillation. T2D: type 2 diabetes. LDL: low-density lipoprotein. AoU: the *All of Us* research program. UKB: UK Biobank. N carriers: number of participants who carry at least one rare variant within each gene. Results are presented in forest plots, with effect sizes (odds ratios [OR] for AF and T2D, betas for height and LDL) and 95% confidence intervals (95% CI). ORs were plotted on a logarithmic scale.

Our study should be interpreted in the context of the design. First, we analyzed samples from all ancestry groups together, which may not fully address population stratification and thus may result in inflated test statistics. However, the whole-genome regression approach has been shown to account for population structure and relatedness and is an established method for analyzing genetic data from diverse populations.^10,30^ Also, the genomic inflation factors and LDSC intercepts reported in the present study did not indicate inflated test statistics. Second, the disease phenotypes (AF, CAD, and T2D) were defined using electronic health records and self-reported data only, which may result in misclassification and thus limit statistical power.

However, the phenotyping algorithms have been validated in previous studies, and the genetic correlation estimates between our GWAS and corresponding previously published large-scale GWAS indicated good consistency. Third, we only included high-confidence loss-of-function variants in the rare variant analysis, which may not fully represent the associations between genes and phenotypes since other rare variants (e.g., deleterious missense variants) that we did not include may have an impact on phenotypic expression.

In conclusion, we replicated known genetic associations in the current release of the *All of Us* research program, indicating that the dataset is a rich and robust resource for common and rare variant genetic discovery. The results of our analyses support the validity of genetic discovery in this multi-ancestry sample. As more data become available in the coming releases, the use of this dataset will facilitate the advancement of biomedical research in diverse human populations.

## Data Availability

Access to individual-level data in the All of Us research program is available to researchers whose institution has signed a data use agreement with All of Us (https://www.researchallofus.org/register/). All of Us provides a publicly available data browser (https://databrowser.researchallofus.org/) containing aggregate-level participant data for users to explore the available data, including genomic variants. Electronic health records (EHR) data, used for phenotyping, belongs to the registered tier dataset. Whole-genome sequencing data belongs to the controlled tier dataset, which requires additional training to access.

## Data Availability

Access to individual-level data in the *All of Us* research program is available to researchers whose institution has signed a data use agreement with *All of Us* (https://www.researchallofus.org/register/). *All of Us* provides a publicly available data browser (https://databrowser.researchallofus.org/) containing aggregate-level participant data for users to explore the available data, including genomic variants. Electronic health records (EHR) data, used for phenotyping, belongs to the registered tier dataset. Whole-genome sequencing data belongs to the controlled tier dataset, which requires additional training to access.

## Author Contributions

X.W. and S.H.C. conceptualized the study and analyzed the data. S.H.C., S.A.L., and P.T.E. supervised this work. J.R. helped with analysis and manuscript editing. J.K. helped with phenotype definitions. A.R. and K.R.M are members of the *All of Us* research program and provided support for this work, including manuscript review. The *All of Us* Research Program provided all the data used in the current study. H.C., N.S.V., L.O., and G.A.T. provided feedback for this project. X.W., S.H.C., and S.A.L. wrote the manuscript. All co-authors reviewed the manuscript.

## Acknowledgements

We would like to thank the *All of Us* research program participants, as this study and the database are possible because of their contributions. *All of Us* established core values and responsible strategies to sustain public trust in biomedical research. We hope the partnership between the participants and the program will benefit the participants and improve the health of future generations.

