## Supplementary Materials for "Common and Rare Variants Associated with Cardiometabolic Traits across 98,622 Whole-Genome Sequences in the *All of Us* Research Program"

#### Supplemental Materials

**Supplementary Author Information** 2

**Supplementary Figures** 4

Figure S1. Manhattan plot of genome-wide association study of coronary artery disease 4

Figure S2. LocusZoom plot of significant risk locus in the atrial fibrillation GWAS 5

Figure S3. LocusZoom plot of significant risk loci in the type 2 diabetes GWAS 6

Figure S4. LocusZoom plot of significant loci in the low-density lipoprotein GWAS 7

**Supplementary Tables** 9

Table S1. Phenotype definition for atrial fibrillation 9

Table S2. Summary statistics of the lead common variants identified in the *All of Us* GWAS and in prior GWAS 11

**Supplementary Methods** 13

Study population 13

Cardiometabolic phenotypes 13

Whole-Genome Sequencing and variant calling 14

Variant-level and sample-level quality control 15

Principal components 15

Common variant analysis and genetic correlation 16

Rare variant analysis using burden test 17

**References** 19

### Supplementary Author Information

**Flagship Manuscript Writing Group:** Alexander Bick MD, PhD[1], Ginger A Metcalf BS[9], Ashley Able PhD[2], Kelsey R Mayo PhD[2], Lee Lichtenstein MS[5], Shimon Rura BA[7], Robert Carroll PhD[3], Anjene Musick PhD, MPH[15]

**All of Us Genomic PIs:** Eric Boerwinkle PhD[9], Mine S Cicek PhD[13], Kimberly F Doheny PhD[10], Evan E Eichler PhD[12], Stacey Gabriel PhD[8], Richard A Gibbs PhD[9], David Glazer BS[6], Paul Harris PhD[3], Gail P Jarvik MD, PhD[11], Anthony Philippakis MD, PhD[5], Heidi L Rehm PhD[8], Dan Roden MD[4], Stephen N Thibodeau PhD[13], Scott Topper PhD[6]

**Biobank, Mayo:** Ashley L Blegen[14], Samantha J Wirkus[14], Jeffrey G Meyer[14], Mine S Cicek PhD[13], Stephen N Thibodeau PhD[13],

**Data and Research Center, Broad Institute of MIT & Harvard:** Lee Lichtenstein MS[5], Sophie Schwartz PhD[5], M. Morgan Taylor PhD[5], Kristian Cibulskis[5], Andrea Haessly MS[5], Rebecca Asch BLA[5], Aurora Cremer[5], Kylee Degatano MS[5], Akum Shergill[6], Laura Gauthier PhD[6], Eric Banks PhD[5], Anthony Philippakis MD, PhD[5]

**Data and Research Center, Vanderbilt University Medical Center:** Melissa Basford MBA[2], Alexander Bick MD, PhD[1], Ashley Able PhD[2], Kelsey R Mayo PhD[2], Robert Carroll PhD[3], Jennifer Zhang MS[2], Henry Condon[1], Yuanyuan Wang PhD[2], Paul Harris PhD[3], Dan Roden MD[4]

**Data and Research Center, Verily:** Shimon Rura BA[7], Moira K Dillon BS[7], CH Albach BS[7], David Glazer BS[6]

**Genome Center, Baylor College of Medicine:** Richard A Gibbs PhD[9], Eric Boerwinkle PhD[9], Donna M Muzny MS[9], Ginger A Metcalf BS[9], Eric Venner PhD[9], Kimberly Walker MS[9], Jianhong Hu PhD[9], Harsha Doddapaneni PhD[9], Christie L Kovar BS[9], Mullai Murugan BS[9], Shannon Dugan BS[9], Ziad Khan BS[9]

**Genome Center, Broad Institute of MIT & Harvard:** Stacey Gabriel PhD[8], Heidi L Rehm PhD[8], Scott Topper PhD[6], Niall J Lennon PhD[8], Namrata Gupta PhD[8], Alicia Zhou PhD[6], Cynthia Neben PhD[6], Christopher Kachulis PhD[8]

**Genome Center, Johns Hopkins University School of Medicine:** Kimberly F Doheny PhD[10], Michelle Z Mawhinney MS[10], Sean ML Griffith MS[10], Elvin Hsu BS[10], Hua Ling PhD[10], Marcia K Adams MS[10]

**Genome Center, University of Washington School of Medicine:** Gail P Jarvik MD, PhD[11], Evan E Eichler PhD[12], Joshua D Smith MS[12], Christian D Frazar MS[12], Colleen P Davis BS[12], Karynne E Patterson BS[12], Marsha M Wheeler PhD[12], Sean McGee PhD[12], Aparna Radhakrishnan PhD[12]

**NIH *All of Us* Research Program Staff:** Andrea H Ramirez MD, MS[15], Sokny Lim MPH[15], Siddhartha Nambiar PhD[15], Anjene Musick PhD, MPH[15], Bradley Ozenberger PhD[15], Chris Lunt BS[15], Joshua Denny MD, MS[15]

[1]Department of Genetic Medicine, Vanderbilt University Medical Center, Nashville, TN, 37203, USA, [2]Vanderbilt Institute of Clinical and Translational Research, Vanderbilt University Medical Center, Nashville, TN, 37203, USA, [3]Department of Biomedical Informatics, Vanderbilt University Medical Center, Nashville, TN, 37203, USA, [4]Department of Clinical Pharmacology, Vanderbilt University Medical Center, Nashville, TN, 37203, USA, [5]Broad Institute of MIT and Harvard, Data Sciences Platform, Cambridge, MA, 02142, USA, [6] Broad Institute of MIT and Harvard, Data Sciences Platform, Cambridge, MA, 02142, USA, [7]Verily Life Sciences, South San Francisco, CA, USA, 94080, [8]Broad Institute or MIT and Harvard, Cambridge, MA 02142 USA, [9]Human Genome Sequencing Center, Baylor College of Medicine, Houston, TX , 77030 USA, [10]Deparment of Genetic Medicine, Johns Hopkins University School of Medicine, Baltimore, MD 21205, USA, [11]Department of Medicine, Division of Medical Genetics, University of Washington School of Medicine, Seattle, WA 98195 USA, [12]Department of Genome Sciences, University of Washington School of Medicine, Seattle, WA 98195, USA, [13]Department of Laboratory Medicine and Pathology, Mayo Clinic, Rochester, MN 55905, USA, [14]Center for Individualized Medicine, Biorepository Program, Mayo Clinic, Rochester, MN 55905, USA, [15]All of Us Research Program, National Institutes of Health, Bethesda, MD 20817

### Supplementary Figures

#### Figure S1. Manhattan plot of genome-wide association study of coronary artery disease

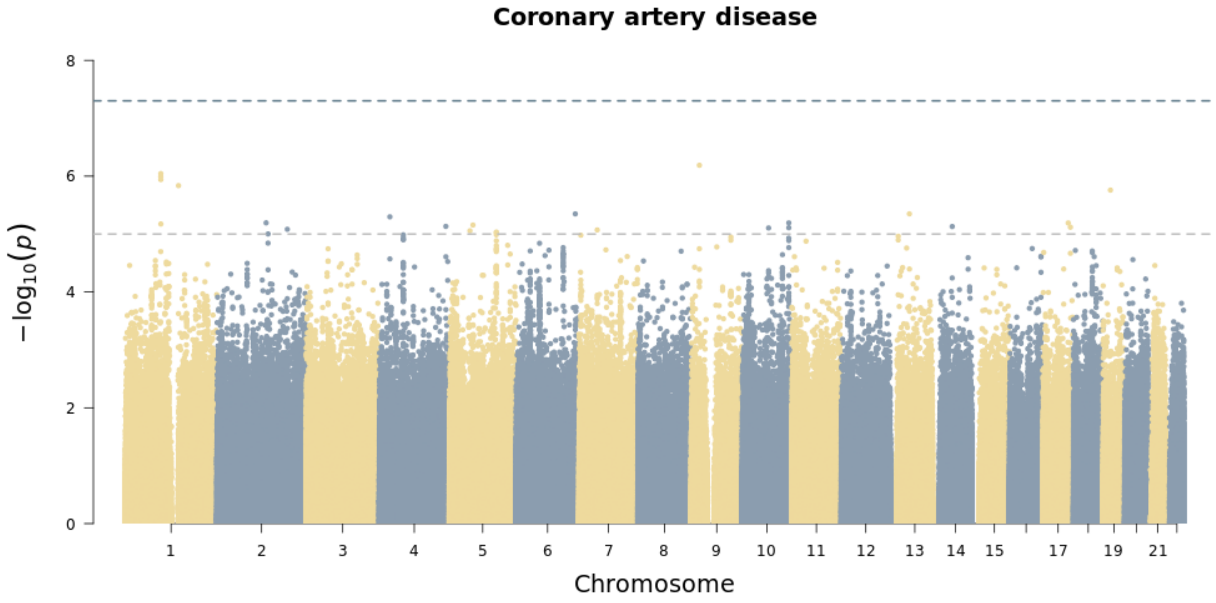


Chromosomal variant positions are plotted on the x-axis. The -log10(*P* values) are plotted on the y-axis. The genome-wide significance threshold ($5\times10^{-8}$) is indicated by the horizontal dotted line.

Figure S2. LocusZoom plot of significant risk locus in the atrial fibrillation GWAS

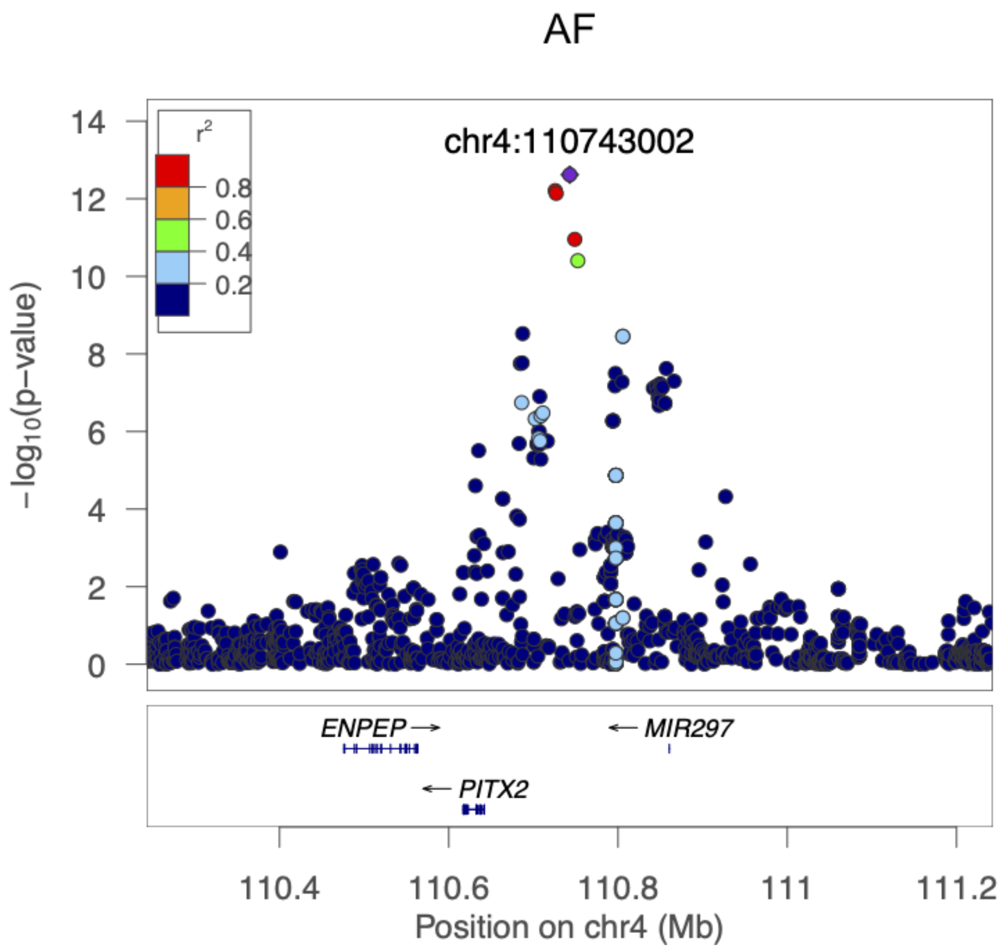

Figure S3. LocusZoom plot of significant risk loci in the type 2 diabetes GWAS

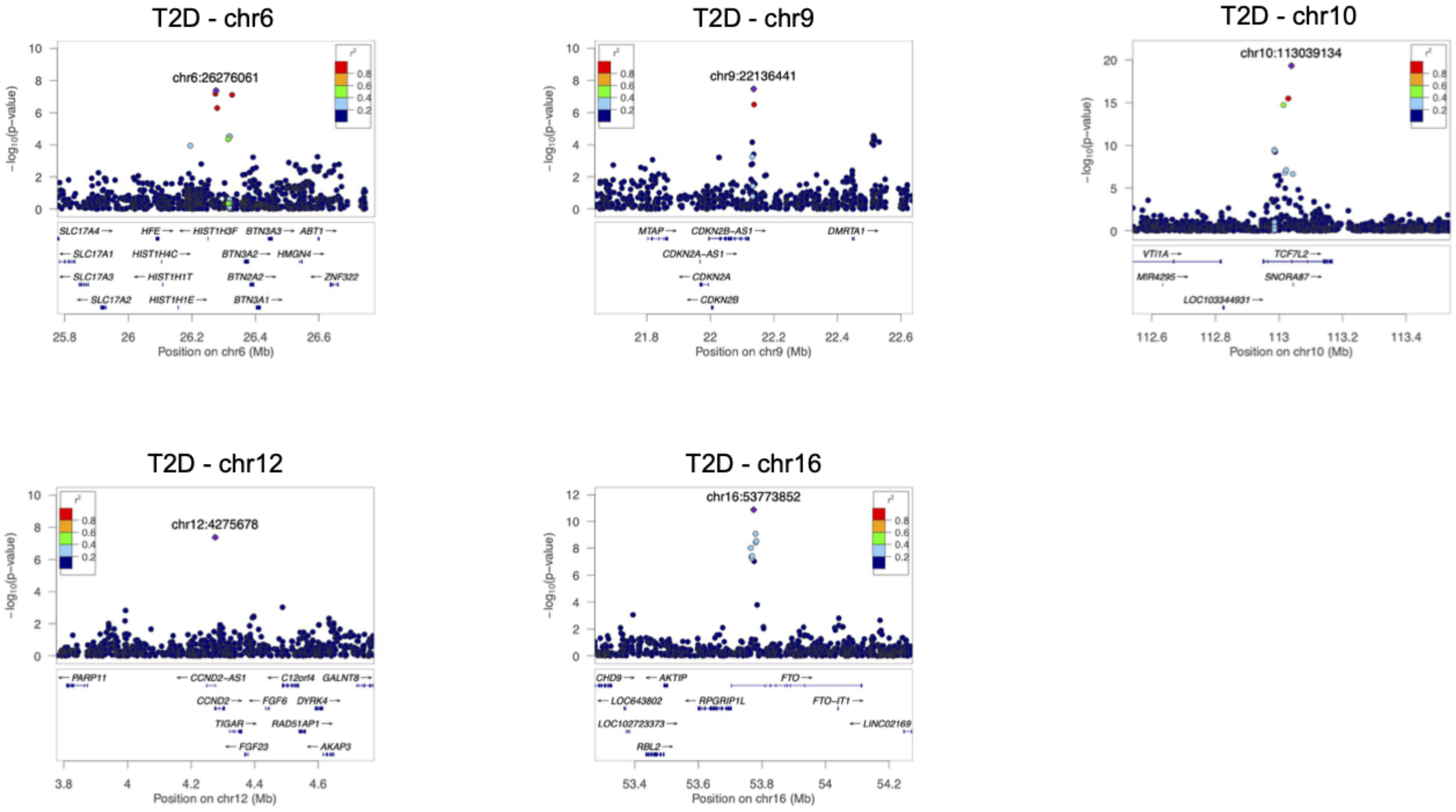

Figure S4. LocusZoom plot of significant loci in the low-density lipoprotein GWAS

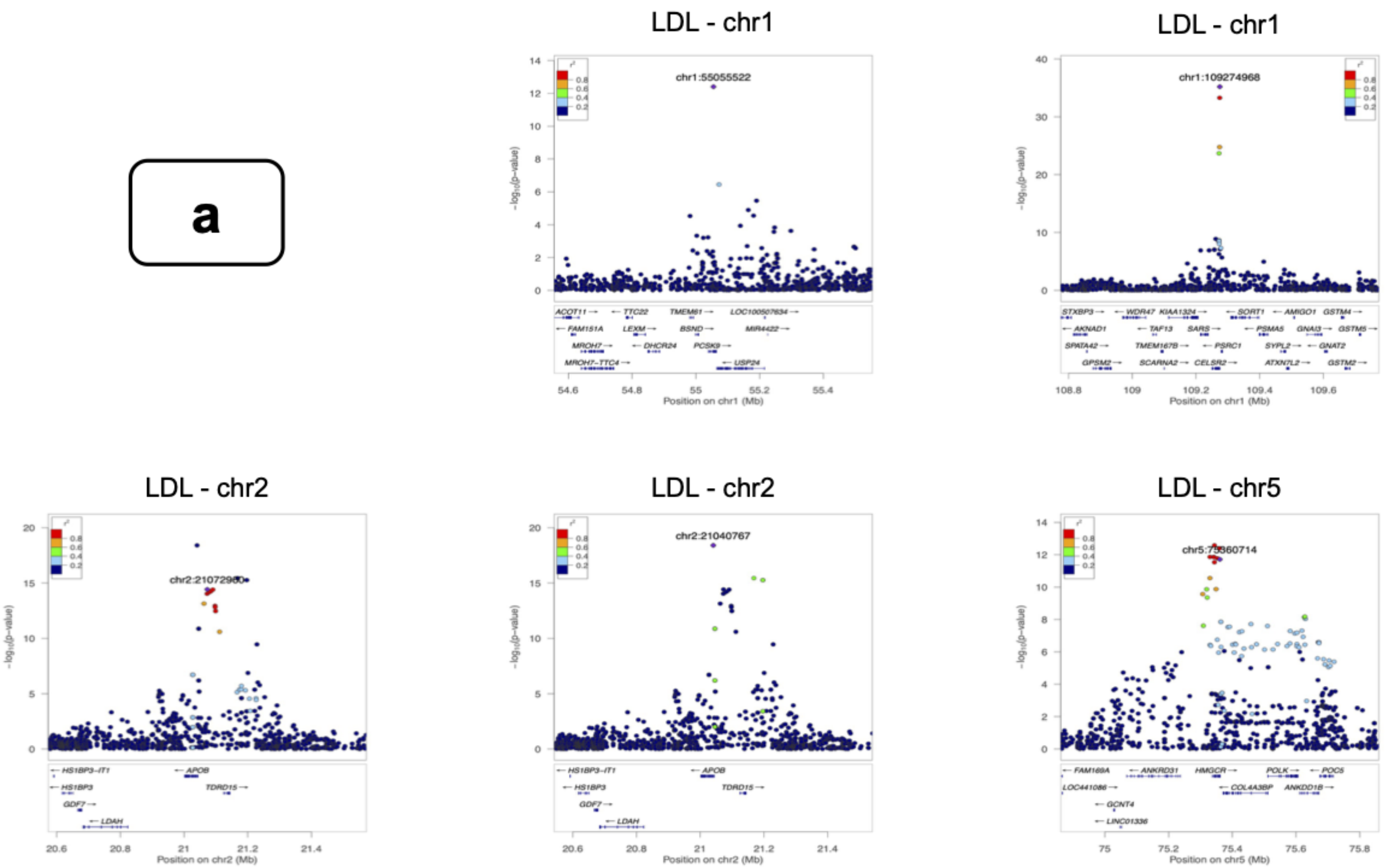

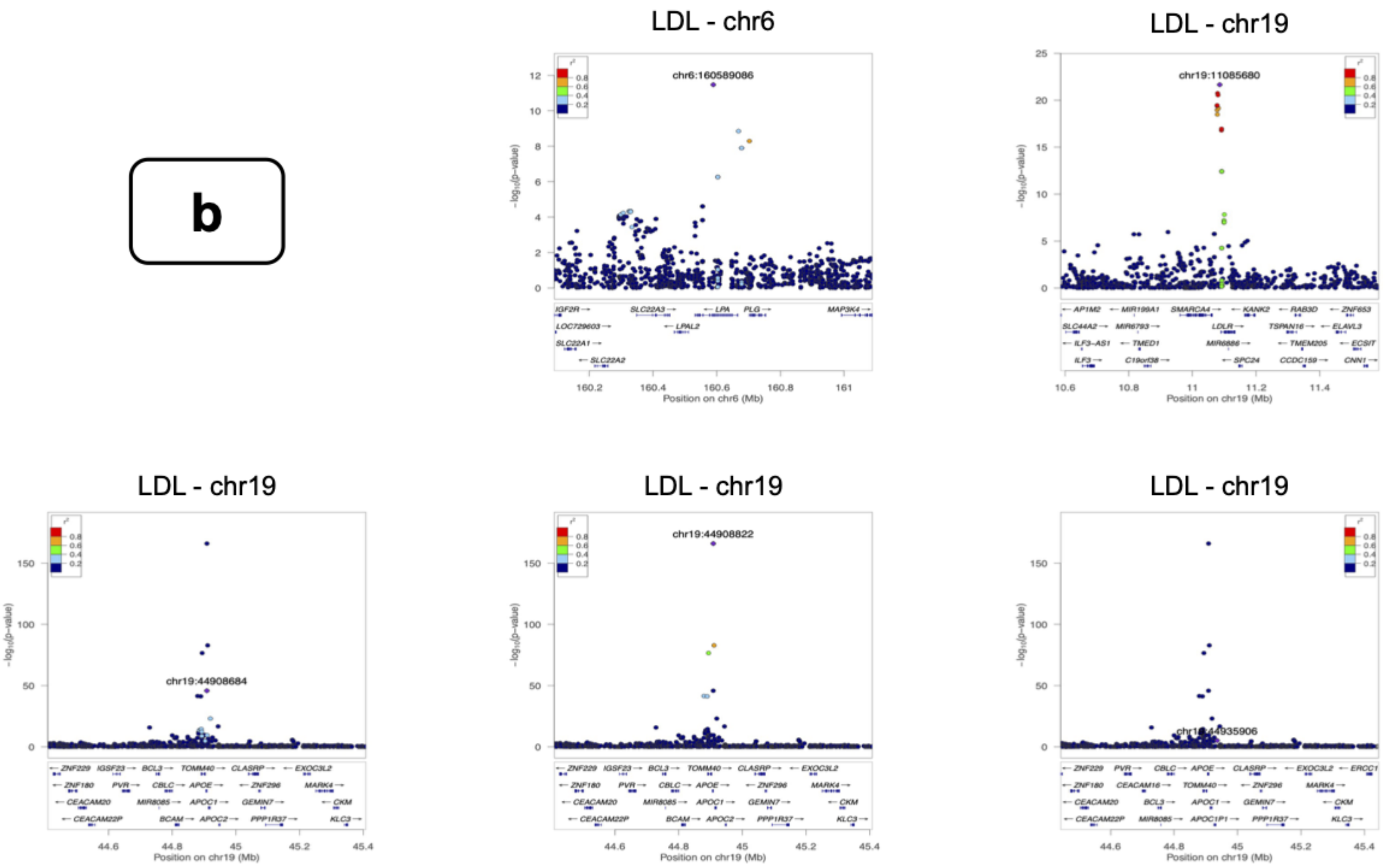
b
LDL - chr6
chr6:160589086
Position on chr6 (Mb)
LDL - chr19
chr19:11085680
Position on chr19 (Mb)
LDL - chr19
chr19:44908684
Position on chr19 (Mb)
LDL - chr19
chr19:44908822
Position on chr19 (Mb)
LDL - chr19
chr19:44935906
Position on chr19 (Mb)

### Supplementary Tables

#### Table S1. Phenotype definition for atrial fibrillation

| | |
|---|---|
| ICD codes | ICD 9: 427.3, 427.31, 427.32<br>ICD 10: I48, I48.0, I48.1, I48.2, I48.3, I48.4, I48.9 |
| Self-reported medical history | Circulatory Conditions: Atrial Fibrillation (Concept id: 43528442) |
| Procedures/Operations | 1. Cardioversion, elective, electrical conversion of arrhythmia; external (CPT4 Code: 92960; Concept Id: 2313791)<br>2. Cardioversion (SNOMED Code: 250980009; Concept Id: 4353741)<br>3. Atrial cardioversion (SNOMED Code: 26879000; Concept Id: 4098410)<br>4. External electrode cardioversion (SNOMED Code: 275148007; Concept Id: 4166447)<br>5. Cardioversion, elective, electrical conversion of arrhythmia; internal (separate procedure) (CPT4 Code: 92961, Concept Id: 2313792)<br>6. Comprehensive electrophysiologic evaluation including transseptal catheterizations, insertion and repositioning of multiple electrode catheters with induction or attempted induction of an arrhythmia including left or right atrial pacing/recording when nec |

| | |
|---|---|
| | (CPT4 Code: 93656; Concept Id: 43528008) |

Concept IDs are codes specific to the *All of Us* database and can be used to query data. Participants with any of these codes in their electronic health records (EHR) were identified as cases for atrial fibrillation.

Table S2. Summary statistics of the lead common variants identified in the *All of Us* GWAS and in prior GWAS

| Genetic variants in the AoU GWAS [ref/effect] (GRCh38) | Effect size [95% CI] in the AoU GWAS | *P*-value in the AoU GWAS | Genetic variants in prior GWAS [ref/effect] (GRCh37) | Effect size [95% CI] in prior GWAS | *P*-value in the GWAS |
|---|---|---|---|---|---|
| **Atrial Fibrillation** | | | | | |
| chr4:110743002 [G/A] | 1.26 [1.18, 1.34] | $2.40\times10^{-13}$ | chr4:111664158 [G/A] | 1.56 [1.53, 1.59] | $8.80\times10^{-453}$ |
| **Type 2 Diabetes** | | | | | |
| chr6:26276061 [T/G] | 0.79 [0.73, 0.86] | $4.22\times10^{-8}$ | Not in prior GWAS | | |
| chr9:22136441 [G/C] | 1.11 [1.07, 1.16] | $3.39\times10^{-8}$ | Not in prior GWAS | | |
| chr10:113039134 [T/A] | 1.19 [1.15, 1.24] | $4.87\times10^{-20}$ | Not in prior GWAS | | |
| chr12:4275678 [T/G] | 0.66 [0.57, 0.77] | $4.23\times10^{-8}$ | chr12:4384844 [T/G] | 0.66 [0.63, 0.68] | $1.20\times10^{-96}$ |
| chr16:53773852 [A/G] | 1.12 [1.08, 1.16] | $1.33\times10^{-11}$ | chr16:53807764 [A/G] | 1.08 [1.05, 1.10] | $2.70\times10^{-9}$ |
| **Low-density lipoprotein** | | | | | |
| chr1:55055522 [C/T] | -0.09 [-0.11, -0.06] | $1.71\times10^{-12}$ | chr1:55521195 [C/T] | -0.08 [-0.06, -0.09] | $3.70\times10^{-20}$ |

| | | | | | |
|---|---|---|---|---|---|
| chr1:109274968 [G/T] | -0.12 [-0.13, -0.10] | $3.49\times10^{-38}$ | chr1:109817590 [G/T] | -0.12 [-0.12, -0.11] | $9.58\times10^{-411}$ |
| chr2:21040767 [T/G] | 0.10 [0.08, 0.12] | $1.59\times10^{-21}$ | chr2:21263639 [T/G] | 0.10 [0.09, 0.11] | $3.53\times10^{-195}$ |
| chr2:21072960 [G/A] | 0.07 [0.06, 0.09] | $7.75\times10^{-20}$ | chr2:21295832 [G/A] | 0.06 [0.06, 0.07] | $1.34\times10^{-174}$ |
| chr5:75360714 [T/C] | 0.06 [0.04, 0.07] | $1.18\times10^{-12}$ | chr5:74656539 [T/C] | 0.06 [0.06, 0.06] | $5.46\times10^{-173}$ |
| chr6:160589086 [A/G] | 0.12 [0.09, 0.16] | $2.94\times10^{-12}$ | chr6:161010118 [A/G] | 0.08 [0.07, 0.09] | $4.80\times10^{-79}$ |
| chr19:11085680 [AC/A] | -0.13 [-0.15, -0.10] | $3.73\times10^{-27}$ | Not in prior GWAS | | |
| chr19:44908684 [T/C] | 0.18 [0.16, 0.20] | $9.51\times10^{-62}$ | chr19:45411941 [T/C] | 0.18 [0.18, 0.19] | $9.12\times10^{-828}$ |
| chr19:44908822 [C/T] | -0.40 [-0.42, -0.37] | $2.03\times10^{-181}$ | chr19:45412079 [C/T] | -0.48 [-0.48, -0.47] | $2.91\times10^{-3040}$ |
| chr19:44935906 [C/G] | -0.03 [-0.05, -0.02] | $1.46\times10^{-4}$ | chr19:45439163 [C/G] | -0.02 [-0.03, -0.01] | $1.96\times10^{-10}$ |

The summary statistics in prior GWAS were extracted from the GWAS used in genetic correlation analysis. When the SNV was not available in the reference GWAS, the summary statistics were obtained from the GWAS catalog. SNVs were noted as not available if it has not been reported in the GWAS catalog. Effect size: odds ratios (OR) for disease phenotypes, beta for continuous traits.

### Supplementary Methods

#### Study population

One of the goals set by the *All of Us* research program was to recruit individuals that have been and continue to be underrepresented in biomedical research due to limited access to health care. Therefore, *All of Us* takes demographics, including race, ethnic group, age, sex, gender identity, income, educational attainment, and geographic location, into account when enrolling participants.[1] For the first release of the genomic data, *All of Us* prioritized historically underrepresented individuals in the sequencing procedure, resulting in a 51% percentage of participants in racial and ethnic minorities among the 98,622 sequenced individuals. The detailed ancestry summary statistics (both genetically predicted and self-reported) are presented in **Table 1**. All participants completed electronic consent modules and health questionnaires upon enrollment, and the study protocol has been published previously.[2] In the current release, all samples with genetic data have at least one other type of data that can be used for research purposes. Approval to use the dataset for the specified demonstration projects was obtained from the *All of Us* Institutional Review Board.

#### Cardiometabolic phenotypes

The cardiometabolic traits included in the present study were atrial fibrillation (AF), coronary artery disease (CAD), type 2 diabetes (T2D), body height, and low-density lipoprotein (LDL). AF was defined using a combination of International Classification of Diseases (ICD) codes, self-reported personal medical history, and procedure and operation codes. The detailed algorithm for AF is described in **Table S2**. For CAD and T2D, we used published phenotype algorithms obtained from the eMERGE network to define the disease phenotypes using electronic health records (EHR) data.[3,4] The CAD algorithm was based on ICD and CPT codes, and the T2D algorithm used information from ICD codes, medication use, and laboratory test results. Body height was extracted from the program's physical measurements data section, which includes data for blood pressure, height, weight, waist circumference, hip circumference, and heart rate, all were measured at enrollment by the program. LDL cholesterol levels were extracted from EHR with the unit of mg/dL. When there were multiple laboratory results for LDL in a participant's EHR, the most recent record was used. Rank-based inverse normal transformation was applied to continuous traits before association testing.

#### Whole-Genome Sequencing and variant calling

Each Genome Center performed quality control (QC) of the specimens obtained from the *All of Us* Biobank. Sample preparation and normalization and DNA library construction have been reported previously.[5] The Illumina NovaSeq 6000 instrument was used to conduct the whole-genome sequencing (WGS) procedure following the manufacturer's best practices. Post-sequencing analysis was performed using Illumina's DRAGEN pipeline, which was harmonized (v3.4.12) between different Genome Centers. The GRCh38 reference genome was used in the alignment step. The single sample QC processes checked fingerprint concordance (array vs. WGS data), sex concordance (genetically determined vs. self-reported), cross-individual contamination rate and coverage to detect major errors, such as sample swaps or contamination. Participants who failed these tests were removed from the current release. The WGS variants were called jointly to reduce systematic biases. Additional sample QC procedures were then performed on the joint callsets, including hard threshold flagging (e.g., number of SNPs: < 2.4M and > 5.0M) and population outlier flagging. Variants QC was performed after sample QC, flagging specific variants from a callset. Processes included hard threshold filters (e.g., ExcessHet, QUAL score) and Allele-Specific VariantQualityScoreRecalibration (AS-VQSR or VQSR, a machine learning technique for identifying variants that are likely artifacts).

#### Variant-level and sample-level quality control

In addition to the QC procedures performed by the program when producing the genomic data, we applied several variant-level and sample-level filterings to keep only high-quality data in the present analysis. For genetic variants that passed internal QC, we further filtered out monomorphic variants, variants in low-complexity regions, variants with call rate < 95%, and variants with Hardy-Weinberg equilibrium $P$ value $< 1 \times 10^{-15}$. We conducted sample QC by excluding samples with call rate < 95%, Ti/Tv ratio, het/hom ratio, SNP/Indel ratio, or number of singletons > 8 standard deviation (SD) from the population mean. We performed the QC procedures using PLINK 2.0 [https://www.coggenomics.org/plink/2.0/].

#### Principal components

Population structure inference on the entire study population was obtained by implementing an algorithm (PC-AiR) that accounts for relatedness in the sample.[6] This method takes the kinship inference obtained using the KING software[7] as input, which assigns negative estimates to pairs of individuals with different ancestry backgrounds. PC-AiR uses these negative kinship estimates to identify groups of participants with different ancestry backgrounds and performs principal component analysis (PCA) on unrelated samples who are representative of the ancestries presented in the entire sample. For the related subset, the algorithm predicts PC values for them based on their genetic similarities with the unrelated subset. Genetic similarities were also estimated using the kinship inference algorithm that accounts for unknown population substructure.[7] We followed a 2-step method presented in the TOPMed analysis pipeline (https://uw-gac.github.io/topmed_workshop_2017/index.html) to calculate the principal components. R package *GENESIS* was used to implement the PC-AiR and PC-Relate algorithms needed for this analysis.

#### Common variant analysis and genetic correlation

We performed association testing for phenotypes and individual genetic variants with minor allele frequencies (MAF) > 1% using a whole-genome regression approach implemented in the REGENIE[8] software assuming an additive genetic model. This method first generates LOCO (leave-one-chromosome-out) predictions of trait values in step 1 using a set of high-quality genetic variants that provide whole-genome information. We used genetic variants with a MAF > 1%, minor allele count (MAC) > 100, missingness rate < 1%, Hardy-Weinberg equilibrium test $P$ value > $1\times10^{-15}$ and linkage-disequilibrium (LD) pruning ($r^2 < 0.1$). The resulting LOCO predictions are then used in step 2 to test the associations between phenotypes and each individual genetic variant. The covariates adjusted in all statistical models were: (1) age (age at enrollment for disease phenotypes, age at measurement for continuous traits), (2) sex, (3) 20 principal components of ancestry. Age at enrollment was calculated using birthdate and program consent date. When consent date was missing from the database, we used median consent date among the samples to calculate the enrollment age. 57,239 participants have consent date information available in the database, ranging from 2017-05-31 to 2021-04-01 (median date: 2019-03-08).

We fitted linear regression models for continuous traits and logistic regression models for binary traits with a saddle point approximation (SPA) method[9] accounting for case-control imbalance. Genome-wide significant variants were considered those with $P < 5\times10^{-8}$. We also conducted conditional analyses[10] using the GCTA software to select secondary independent significant SNVs at each locus. The mapped gene at each locus was either the nearest gene or the phenotype-associated gene within 500 kb range of the lead variant reported in the GWAS catalog. We then estimated genetic correlation using GWAS summary statistics to evaluate how consistent our GWAS results were with corresponding previously published GWAS results. We used the LD score regression approach[11] implemented in the LDSC software and pre-computed LD-scores provided by the software for this analysis. LDSC is not a bounded estimator and thus may generate estimates less than -1 or greater than 1 due to sampling variation. Also, a genetic correlation estimate that is greater than 1 may indicate that the true genetic correlation is high and there are some samples that overlap between the two studies. All correlation estimates for disease phenotypes were converted from observed scale to liability scale using sample prevalence and population prevalence estimates.

#### Rare variant analysis using burden test

To test the association between each phenotype and the burden of rare variants (MAF < 0.1%) of their known associated genes, we first annotated the protein consequences of rare variants using the Loss-of-Function Transcript Effect Estimator (LOFTEE)[12] plug-in implemented in the Variant Effect Predictor105 (VEP; v.95) (https://github.com/konradjk/loftee) software to identify high-confidence loss-of-function (LoF) variants, including frameshift indels, stop-gain variants and splice site disrupting variants. Furthermore, we checked the continental allele frequencies using gnomAD v2 to make sure the maximum population frequency (POPMAX) for each of the rare variations was also < 0.1% since ancestral allele frequencies can be different from the pooled ones. We identified 597 LoF variants within *TTN*, 38 within *GIGYF1*, 32 within *APOB*,

17 within *LDLR*, 14 within *PCSK9*, 49 within *ADAMTS17*, 15 within *ACAN*, and 32 within *NPR2*. We then tested the association between the burden of selected rare variants in each gene and its corresponding reported phenotype using a burden test implemented in the REGENIE software.[8] Specifically, for each gene, we counted the number of alternative allele copies each individual carries and treated this number as a single burden genotype. This genotype was then used in a regression model (linear models for continuous traits, logistic models for binary traits) to associate with the phenotype, adjusting for age (age at enrollment for disease phenotypes, age at measurement for continuous traits), sex, and 20 principal components of ancestry. The LOCO predictions obtained from common variant analysis were used in step 2 for this analysis. We also applied the saddle point approximation (SPA) method[9] to account for case-control imbalance.
